# Neurological manifestations of coronavirus infections – a systematic review

**DOI:** 10.1101/2020.08.26.20182196

**Authors:** Jesper Almqvist, Tobias Granberg, Antonios Tzortzakakis, Stefanos Klironomos, Evangelia Kollia, Claes Öhberg, Roland Martin, Fredrik Piehl, Russell Ouellette, Benjamin V. Ineichen

## Abstract

In order to optimize diagnostic workup of the current severe acute respiratory syndrome coronavirus 2 (SARS-CoV-2) pandemic, we systematically reviewed neurological and neuroradiological manifestations of SARS-CoV-2 and all other known human coronavirus species (HCoV). Which lessons can we learn? We identified relevant publications (until July 26^th^ 2020) using systematic searches in PubMed, Web of Science and Ovid EMBASE with predefined search strings. A total of 4571 unique publications were retrieved, out of which 378 publications were selected for in-depth analysis by two raters, including a total of 17549 (out of which were 14418 SARS-CoV-2) patients. Neurological complications and associated neuroradiological manifestations are prevalent for all HCoVs (HCoV-229E, HKU1, NL63, OC43, Middle East respiratory syndrome (MERS)-CoV, SARS-CoV-1 and SARS-CoV-2). Moreover, there are similarities in symptomatology across different HCoVs, particularly between SARS-CoV-1 and SARS-CoV-2. Common neurological manifestations include fatigue, headache and smell/taste disorders. Additionally, clinicians need to be attentive for at least five classes of neurological complications: (1) Cerebrovascular disorders including ischemic stroke and macro/micro-hemorrhages, (2) encephalopathies, (3) para-/postinfectious immune-mediated complications such as Guillain–Barré syndrome and acute disseminated encephalomyelitis, (4) (meningo-)encephalitis, potentially with concomitant seizures and (5) neuropsychiatric complications such as psychosis and mood disorders. Our systematic review highlights the need for vigilance regarding neurological complications in patients infected by SARS-CoV-2 and other HCoVs, especially since some complications may result in chronic disability. Neuroimaging protocols should be designed to specifically screen for these complications. Therefore, we propose practical imaging guidelines to facilitate the diagnostic workup and monitoring of patients infected with HCoVs.

## Introduction

Neurological complications of coronavirus disease 2019 (COVID-19), caused by severe acute respiratory syndrome coronavirus 2 (SARS-CoV-2), are emerging. Meanwhile, there are opportunities to learn from previous experiences with other human coronaviruses (HCoVs), among them SARS-CoV-1, triggering an epidemic in 2002,^1^ and Middle East respiratory syndrome coronavirus (MERS-CoV), responsible for an epidemic in 2012.^2^ Moreover, four additional HCoV species (HCoV-229E, HCoV-HKU1, HCoV-NL63 and HCoV-OC43) cause disease in humans, albeit typically with milder clinical courses.^3^

While coronaviruses primarily target the human respiratory system,^4^ they can also enter the central nervous system (CNS). This is evident from pre-clinical research, where murine coronaviruses have been used to model human encephalitis for decades.^5^ HCoVs also show neurotropism. SARS-CoV-1^6^ and SARS-CoV-2^7^ both use the cell membrane-bound human angiotensin-converting enzyme 2 for cellular entry, which is, among other tissues, expressed on vascular endothelial cells in the brain.^8^ Yet, SARS-CoV-2 seems to have an even higher affinity to bind to this receptor compared to SARS-CoV-1.^9^ SARS-CoV-1 and SARS-CoV-2 are also most related genetically, with 79% genome homology.^10^ Therefore, it is not surprising that several studies have described neurological symptomatology in both SARS-CoV-1 and SARS-CoV-2 infections.^3, 11^ Neurological complications have also been described in other HCoVs.^12^

The aim of this study was to systematically summarize neurological and neuroimaging manifestations of all known HCoVs in order to provide possibilities to predict short- and longterm neurological complications of COVID-19. The underlying hypothesis was that there are similarities, in particular between SARS-CoV-1 and SARS-CoV-2, that can help guide our current and future diagnostics of neurological complications to minimize the long-term effects of the current SARS-CoV-2 pandemic.

## Methods

We registered the study protocol in the International prospective register of systematic reviews (PROSPERO, CRD42020183405, https://www.crd.york.ac.uk/PROSPERO/) and used the Preferred Reporting Items for Systematic Reviews and Meta-Analysis (PRISMA) Guidelines for reporting.^13^

### Search strategy

We searched for original studies/abstracts on coronavirus and neurological/neuroradiological manifestations published until July 26th 2020 in PubMed, Web of Science and Ovid EMBASE. The search strings used in each of these databases are described in **Table e-1**. Reviews on the topic were also scrutinized for additional references.

### Eligibility criteria

We included publications that reported on any neurological and/or neuroradiological manifestations related to acute or prior coronavirus infection, i.e. neurological symptoms/complications in coronavirus-infected persons (we included studies reporting on any symptom associated with, but not necessarily specific for neurological diseases, e.g. headache, fatigue), neuropathology findings of coronavirus infections as well as neuroimaging findings associated with coronavirus infections. We excluded animal studies, records with non-English abstracts and reviews.

### Study selection and data extraction

Titles and abstracts of studies were independently screened for their relevance in the web-based application Rayyan (https://rayyan.qcri.org/) by two reviewers (J.A. and B.V.I.) followed by full-text screening.^14^ The same reviewers independently extracted title, authors, publication year, study design, country, number of subjects per group, neurological symptoms/complications, neuropathological findings and/or neuroimaging findings associated to acute or prior coronavirus infection. Any results of associations, including whether analyses were adjusted for confounders, were extracted. Discrepancies in findings were resolved by discussion among assessors, a third reviewer (T.G.) was consulted as needed.

### Quality assessment

The quality of each study with more than 10 included subjects was assessed against pre-defined criteria by the same two reviewers using the Newcastle-Ottawa scale for evaluating risk of bias in non-randomized studies and are further described in **Table e-2.^15^**

### Data synthesis and analysis

Studies and results were qualitatively compared and summarized. We did not assess publication bias.

## Results

### Eligible publications

In total, 12195 original publications were retrieved from our comprehensive database search. An additional 13 publications were retrieved by screening relevant reviews. After abstract and title screening, 611 studies were eligible for full-text search of which 371 (8.3% of deduplicated references) and an additional 7 abstract for which the English full-text was not available were included in the qualitative synthesis (**Figure 1**). All references are listed in the supplementary reference list.

**Figure 1:**
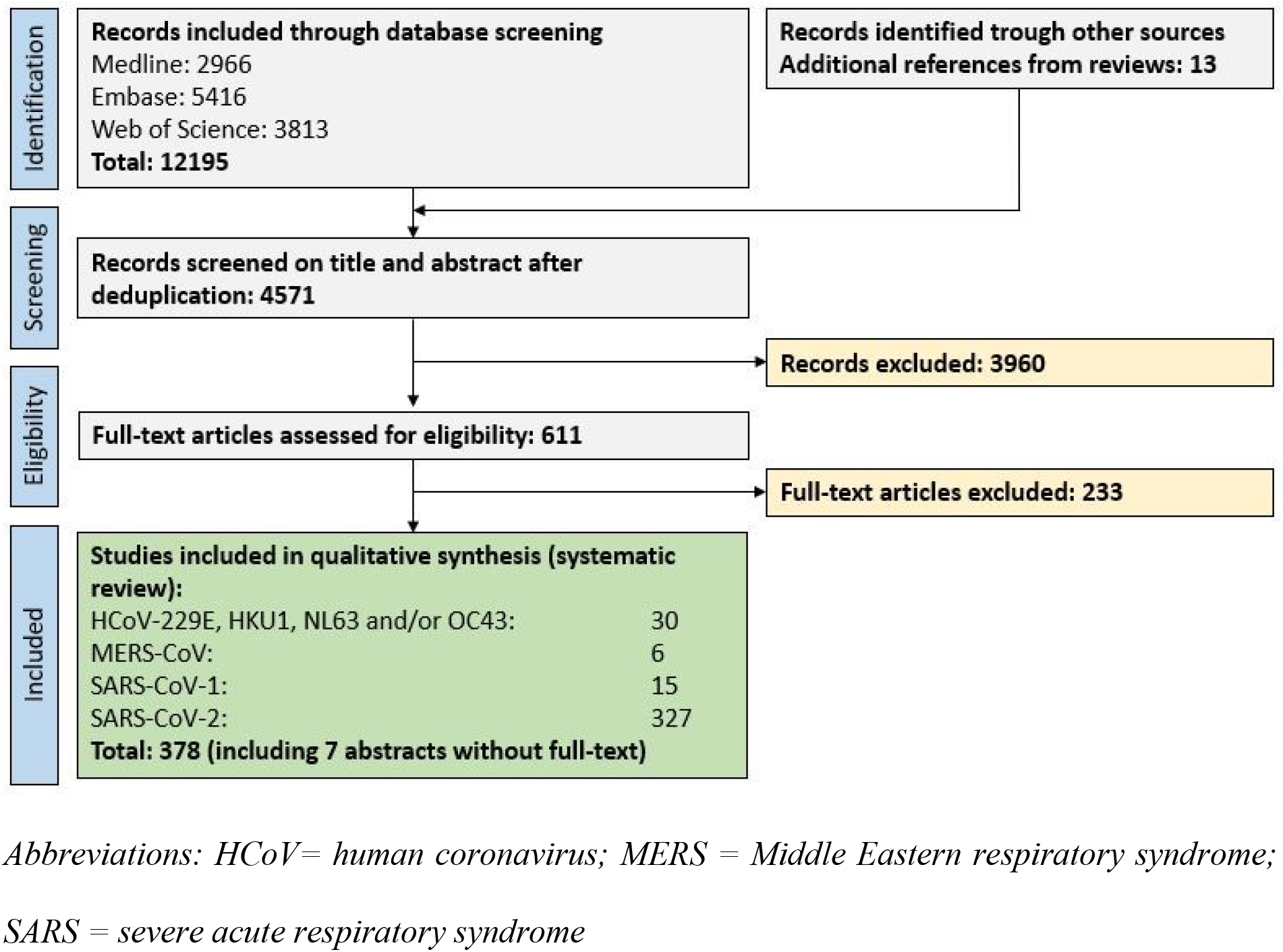
PRISMA flow chart depicting the study selection process.

### Risk of bias assessment

We evaluated the risk of bias for all studies with more than 10 subjects. Most of the studies showed a low risk of bias for the selection domain, as well as for subunits 2 and 3 from the exposure domain, i.e. if patients and controls were treated equally and if studies showed low exclusion rates, respectively (see **Table e-3)**. Many studies did not report on adjusting their statistical analyses for subject age, sex or other potential confounders (comparability domain), thus potentially introducing biases. Of note, a considerable proportion of studies was published as letters or editorials thus hampering a thorough risk of bias assessment.

### General study characteristics

Of the eligible 378 publications, 30 publications investigated clinically more benign strains of HCoV, including a total of 961 subjects. The majority of these publications assessed HCoV OC43 (26 publications), followed by 229E (22), NL63 (7) and NKU1 (5). 6 publications assessed MERS-CoV, comprising a total of 341 patients, while 15 publications assessed SARS-CoV-1, including a total of 1829 cases. 327 publications assessed SARS-CoV-2, with the majority being case reports (179). These studies included a total of 14418 SARS-CoV-2 patients (including 45 children) from 35 different countries. Study characteristics and main findings for every HCoV strain are summarized in **Table e-4 – Table e-7**.

### Association of coronavirus infections with neurological symptoms and complications

#### Clinically milder infections with HCoVs: 229E, HKU1, NL63 and OC43

##### Acute infections

Findings from studies regarding the more clinically benign HCoVs are summarized in **Table e- 4**. Most studies assessed neurological manifestations of coronavirus infections in children. A retrospective cohort study from the USA investigated 1683 respiratory specimens from children and found that 84 of them (5%) were positive for coronavirus (229E, HKU1, NL63 and/or OC43); of these, 5 children (8%) had meningoencephalitis and/or febrile seizures.^16^ A prospective multicenter study from China confirmed the association of coronavirus, particularly HKU1, with febrile seizures: 87 of 4181 consecutive patients (2.1%) were tested positive for coronavirus (229E, HKU1, NL63 and/or OC43).^17^ Five out of 13 children (38%) positive for HKU1 had febrile seizures and 1 adult patient with OC43 had an aseptic meningitis. Two prospective studies from Slovenia investigating the same patient cohort found that 19 out of 278 children (7%) with febrile seizures were tested positive for coronavirus (mostly OC43: 12/19 patients, 63%).^18, 19^ Another prospective multicenter study from Turkey included 192 children with febrile seizures and found that HCoV OC43 is the single most common, yet still rare, cause for febrile seizures in children under 12 months (6% of children under 12 months with febrile seizures were positive for OC43).^20^ Finally, five case reports in children described the following neurological complications associated with the detection of coronavirus: acute flaccid paralysis, acute disseminated encephalomyelitis (ADEM), Guillain–Barré syndrome (GBS) and two cases of fatal encephalitis (both in children with underlying immune deficiency), described in **Table e-4**.

One retrospective cohort study from Finland assessed neurological complications associated with HCoV infections in adults. OC43 was detected in 28 out of 14000 sera from patients with infectious diseases.^21^ Four of these patients suffered either meningitis, convulsions, headache or vertigo.

##### Association of prior infections to neurological/psychiatric diseases

Findings are summarized in **Table e-8**. An early case-control study from 1980 isolated infectious coronavirus particles from autopsy brain material from 2 MS patients (out of 13 MS patients and 12 controls).^22^ Injection of 10% homogenate of the HCoV positive brain material (in saline solution) led to encephalitis in mice within 3 – 5 days. Several subsequent studies confirmed the presence of OC43 and/or 229E in different tissues including brain, serum and cerebrospinal fluid (CSF).^23-25^ Yet, while some studies observed a higher prevalence of coronavirus antibodies in MS patients versus controls, most studies could not confirm this finding (**Table e-8)**.

Two studies have assessed the association of prior coronavirus infections with psychiatric symptoms. A case-control study that included 257 subjects with major depression or suicide attempt, found that seropositivity for coronavirus strain NL63 (as well as Influenza A and B) was associated with the history of mood disorders.^26^ Another case-control study observed higher immunoglobulin G (IgG) titers towards different coronavirus species in patients with recent onset of psychotic symptoms as compared to controls.^27^

#### MERS-CoV

Findings are summarized in **Table e-5**. Two larger retrospective case series from Saudi Arabia assessed symptomatology among MERS-CoV patients. One multicenter study found that 71% of patients (185/227) reported some form of neurological symptoms.^28^ The most common neurological symptoms were fatigue (108/185, 58%), headache (59, 31%), altered consciousness (53, 29%) and/or focal neurological deficits (10, 5%). Additionally, altered level of consciousness at time of diagnosis was a significant risk factor for death. The other study included 70 patients with confirmed MERS, of which the large majority had clinical symptoms (67/70, 96%).^29^ The neurological symptomatology was similar; fatigue (29/67, 43%), confusion (18, 27%), headache (9, 13%) and/or seizures (6, 9%). Four smaller case series/case reports including a total of 10 patients described neurological complications in MERS-HCoV patients, among them toxic/infectious neuropathy, Bickerstaff brainstem-encephalitis with intracranial hemorrhage, ischemic stroke, critical illness neuropathy, brain stem dysfunction and ADEM. Of note, 8 of these patients had pre-existing medical conditions, including diabetes type 2 and/or cardiovascular disease (**Table e-5**).

#### SARS-CoV-1

Findings are summarized in **Table e-6**. Two cohort studies assessed neurological manifestations of adult SARS-CoV-1 patients. One study comprised 206 SARS-CoV-1 cases, constituting all SARS-CoV-1 patients from Singapore.^30^ 5 of these patients (2.4%) suffered large artery cerebral strokes. It is noteworthy that only one of these patients had pre-existing risk factors for cerebrovascular disease. The other cohort study included 1291 Chinese SARS-CoV-1 patients and investigated how cerebrocardiovascular disease affected SARS-CoV-1 disease outcomes (only abstract available in English).^31^ Critical conditions and multi-organ dysfunction occurred more frequently among SARS-CoV-1 patients with pre-existing cerebrocardiovascular diseases (58% and 27%, respectively) compared to patients without underlying diseases (28% and 10%, respectively). One cross-sectional study aimed at assessing neuropsychiatric symptoms during acute and convalescence phases of SARS-CoV-1 via a questionnaire sent out to 308 patients.^32^ The response rate was 33% (102 patients); 65% of these patients had suffered neuropsychiatric symptoms during the convalescence phase of SARS-CoV-1. Steroid treatment during hospitalization was predictive for anxiety, depression, psychosis and behavioral symptoms during both the acute and convalescence phases. Another Chinese study (only abstract available in English) including 173 SARS-CoV-1 patients confirmed high rates of neurological/neuropsychiatric symptoms in SARS-CoV-1 patients (53%).^33^ The symptomatology comprised headache (67%), affective disorder (31%), dizziness (29%), anxiety (20%), reduced consciousness (10%), phobia (8%), depression (6%), unspecified mental disorder (5%), suicidal ideation (1%), seizures (1%) and focal neurological signs (0.6%). Several case reports, comprising a total of 11 patients, described neurological complications in SARS-CoV-1, among them critical illness neuro-/myopathy, seizures, persistent sleeping difficulties, persistent anosmia, delirium and generalized pain (**Table e-6**).

##### Children

One cohort study assessed neurological manifestation of SARS-CoV-1 in children.^34^ The study comprised 183 children with encephalitis out of which 22 (12%) were positive for SARS-CoV- 1. Headache, seizures and neck stiffness were common symptoms among SARS-CoV-1 positive children with encephalitis (10/22 [46%], 5/22 [23%] and 7/22 [32%], respectively). A total 16 of these children underwent neuroimaging out of which 8 (50%) showed abnormal findings, among them abnormalities in the temporal lobe, in the periventricular region and in the basal ganglia/thalamus. This study also measured different inflammatory proteins in serum and CSF; in matched serum-CSF samples, interleukin (IL)-6, IL-8 and chemokine (C-C motif) ligand 2 were increased in CSF compared to serum.

##### Neuropathology

Several studies addressed neuropathological changes of SARS-CoV-1. One relatively large study including findings from 18 autopsies (out of which 8 were confirmed SARS-CoV-1 infections pre-mortem) detected SARS-CoV-1 infected neurons in the hypothalamus and cortex using in situ hybridization and electron microscopy.^35^ Six of eight patients (75%) showed diffuse hypoxic neuronal injury. Interestingly, SARS-CoV-1 viral sequences and pathologic changes were unique to brains of SARS-CoV-1-confirmed cases. A smaller case series including 3 patients who died from SARS-CoV-1 showed signs of systemic vasculitis engaging smaller brain vessels.^36^ In one case with CT findings suggestive of ischemia, the histopathological exam showed signs of neuronal necrosis and glial cell hyperplasia.^37^ One study including 5 SARS-CoV-1 patients found changes in cell numbers/staining intensities compared to controls in various endocrine cell pools of the adenohypophysis.^38^ Finally, one study found myopathic changes in 4 out of 8 SARS-CoV-1 cases (50%) undergoing autopsy (in situ hybridization for SARS-CoV-1 in the skeletal muscle was negative though).^39^

#### SARS-CoV-2

##### Neurological symptomatology

Findings are summarized in **Table e-7**. Two large retrospective studies from Spain and China including 841 patients and 214 patients reported that 57% and 38% of patients had neurological manifestations, respectively. ^40, 41^ Both studies also found that neurological symptoms were more common in patients with more severe disease courses.

Several common neurological symptoms among SARS-CoV-2 patients have been described in these studies, such as fatigue (44 – 64% of patients),^42-44^ headache (8 – 14% of patients [up to 70% in 1 study]^45^)^40-44, 46, 47^, dizziness (6%)^41^ and confusion (0.9 – 9% of patients).^44, 46^ Smell and/or taste disorders are other symptoms reported by several studies,^45, 48, 49^ having a prevalence of 39 – 88% in SARS-CoV-2 patients,^47, 50^ Two studies also observed that smell/taste disorders are the initial manifestation of SARS-CoV-2 infection in 36%/60% of patients.^41, 50^ A Spanish survey-based study showed that smell disorders can persist for several weeks in about 1/3 of patients.^51^ Also other symptoms may persist; an Italian study followed 143 patients for an average of 60 days after onset of SARS-CoV-2 symptoms, finding that 76 patients (53%) suffered persisting fatigue.^52^

##### Neurological complications

Several studies reported on neurological complications of SARS-CoV-2, mostly in severe or critically-ill patients. A case series from the UK included 43 SARS-CoV-2 patients who had been admitted to a specialized COVID neurology unit.^53^ Based on symptomatology, patients could be classified into four major subgroups: (1) ischemic stroke (8 patients), (2) inflammatory CNS syndromes (para-/postinfectious, 12 patients), (3) peripheral nervous system (PNS) syndromes such as GBS (7 patients) and (4) encephalopathies (10 patients).

Several larger studies have confirmed these classes of neurological complications in SARS-CoV-2. Cerebrovascular diseases were amongst the most commonly reported complications, including ischemic stroke and cerebral hemorrhages.^41, 54-59^ Studies from China, Spain and the US report that 1 – 1.6% of hospitalized patients were affected by cerebrovascular disease.^41, 54, 55, 57, 59^ Yet, cardiovascular risk factors were pre-existent in many of the stroke patients.^60^ Ischemic brain lesions seem to be more common in more severe disease courses: In a Chinese study, 20% of deceased SARS-CoV-2 patients had signs of hypoxic encephalopathy compared to only 1% in patients who recovered.^43^ Cerebral susceptibility abnormalities (i.e. microbleeds/thrombosis) were also described as a relatively common neuroimaging feature, particularly in atypical locations such as in the corpus callosum or juxtacortically.^61-64^ Several cases of cerebral venous thromboses have been observed.^56, 61^

Inflammatory and/or demyelinating syndromes of both the central and the peripheral nervous system have been reported, among them GBS,^53, 56, 65^ ADEM,^53^ Miller-Fisher syndrome and facial nerve palsy (**Table e-7**).

Encephalopathy and/or encephalitis are also reported by an increasing number of studies. An observational case series from France reported on neurological complications in intensive care unit patients: 13/58 of patients (22%) presented with encephalopathic features.^66^ Brain MRI showed that 8/13 patients displayed leptomeningeal enhancement on post-contrast T_1_-weighted and fluid-attenuated inversion recovery (FLAIR) sequences – a sign of leptomeningeal inflammation. Another French study found that 7/26 patients (27%) showed electroencephalogram (EEG)/MRI changes suggestive of encephalopathy.^64^ Also, several cases of acute necrotizing encephalopathy (ANE) or posterior reversible encephalopathy syndrome (PRES) have been described.^67, 68^ A Turkish study showed that 12/27 patients (44%) showed abnormalities on brain MRI with signs suggestive of ischemia or menigoencephalitis.^69^ Of note, only minimal evidence points to direct infection of neurons/glial cells by SARS-CoV-2 viral particles, as shown by mostly negative RT-PCR analyses from CSF (**Table e-7**).^53, 64, 66^ Nevertheless, CSF from a small subset of patients with encephalitic features did test positive for SARS-CoV-2 RNA.^70^, ^71^

Seizures have been reported in SARS-CoV-2 patients; in a Chinese cohort, 4/78 hospitalized patients (0.5%) suffered at least one seizure.^40^ This was mirrored in a Spanish cohort, where 6/841 hospitalized patients (0.7%) had at least one seizure.^41^ However, a study focusing on seizure prevalence among SARS-CoV-2 patients did not observe any acute symptomatic seizures at all, despite seizure-triggering metabolic changes such as electrolyte imbalance in 84/311 patients (27%).^72^

Finally, several recent studies observed psychiatric complications in SARS-CoV-2 patients. An Italian survey-based study observed sleep impairment as the most commonly reported neurological symptom (94/103 patients).^73^ A prospective cross-sectional study from the USA showed that impaired sense of smell was associated with anxiety and depressed mood.^74^ Finally, a UK-wide surveillance study recorded 23 patients with neuropsychiatric disorders, among them new-onset psychosis (10 patients, 43%), neurocognitive syndrome (6 patients, 26%) and affective disorders (4 patients, 17%).^58^

##### Children

In a study from the UK, 4 out of 27 children (15%) suffering multisystem inflammatory syndromes also developed neurological symptoms, including encephalopathy, brainstem/cerebellar signs and reduced reflexes. MRI showed signal changes of the splenium in all patients, interpreted as a sign suggestive of an ongoing (para-)infectious process.^75^ However, virus was undetectable in CSF from 2/2 sampled children. All patients showed at least some degree of recovery by the end of the study. One case series from China included 8 children with severe SARS-CoV-2 courses:^76^ Of them, one child presented with headache and another with fatigue. Furthermore, cases of children with GBS, stroke and encephalitis have been described (**Table e-7**).

##### Fluid biomarkers

In a Chinese study, patients with neurological symptoms had lower lymphocyte levels, platelet counts as well as higher blood urea nitrogen levels compared with those without neurological symptoms.^40^ Blood neurofilament light chain (NfL) levels have been linked to SARS-CoV-2: SARS-CoV-2 status seems to be an independent predictor of serum NfL levels. Additionally, in severe SARS-CoV-2 patients, an early peak in plasma GFAP was followed by a slower increase in NfL, possibly reflecting a more acute involvement of astrocytes followed by a more prolonged neuro-axonal degeneration.

##### Neuropathology

Several studies assessed neuropathological changes associated with SARS-CoV-2 infections: One case series including 6 SARS-CoV-2 patients found lymphocytic panencephalitis and meningitis and, in 3 patients aged < 65 years, widespread petechial hemorrhages.^77^ Interestingly, all patients also showed signs of brain stem encephalitis. Signs of perivenular inflammation/demyelination suggestive of ADEM have also been described. ^53, 78^ A neuropathological assessment of 18 SARS-CoV-2 patients showed changes consistent with hypoxic injury. However, RT-PCR and immunohistochemistry disclosed only minimal evidence of SARS- CoV-2 infection.^79^ Yet, one SARS-CoV-2 case in a patient with Parkinson’s disease showed electron dense particles suggestive of SARS-CoV-2 in both brain endothelial cells and neurons. SARS-CoV-2 RT-PCR was positive in brain tissue but not in the CSF.^80^

### Comparison between SARS-CoV-1 and SARS-CoV-2

Direct comparison of neurological manifestations between SARS-CoV-1 and SARS-CoV-2 shows substantial similarities (**Table 1)**. In both diseases, a considerable number of patients suffer neurological complications, in particular those with a more severe disease course. Fatigue, headache, smell/taste disorders and reduced consciousness are among the most commonly reported symptoms. For example, the SARS-CoV-2 patient shown in **Figure 2**, presented with altered consciousness to the intensive care unit. Ischemic/hypoxic pathology, including stroke as well as seizures, have been described in both diseases. Similarly, neuropsychiatric complications such as psychosis and mood disorders have been reported for both viral infections. Finally, immune-mediated complications such as GBS, Miller-Fisher syndrome or polyneuritis cranialis have been described in both diseases.

**Table 1.** Comparison between neurological manifestations in SARS-CoV-1 versus SARS- CoV-2, ordered according to evidence level.

| Evidence level | SARS-CoV-1 | SARS-CoV-2 |
| --- | --- | --- |
| **Prospective or retrospective cohort studies** | **<u>Symptoms:</u>**<br>•Children: headache<br>**<u>Complications:</u>**<br>•Cerebrovascular disease<br>•Children: seizures<br>•Pre-existing cerebrocardiovascular disease as risk factor for more severe disease courses | **<u>Symptoms:</u>**<br>•Fatigue (can persist after COVID-19)<br>•Headache<br>•Confusion<br>•Smell/taste disorders (can be persistent)<br>•Dizziness<br>•Neuropsychiatric symptoms including mood disorders and psychosis<br>**<u>Complications:</u>**<br>•Cerebrovascular disease (ischemic stroke/intracranial hemorrhage, microbleeds, cerebral venous thrombosis). Worse ischemic stroke outcome in COVID-19 patients compared to non-COVID-19 patients.<br>•Encephalitis<br>•4% of cases deceased due to neurological complications. |
| **Case-control studies, large case series** | **<u>Symptoms:</u>**<br>•53% of patients with neuropsychiatric symptoms.<br>•Neuropsychiatric symptoms more common in patients with severe disease course<br>•Headache<br>•Affective disorders<br>•Dizziness<br>•Anxiety<br>**<u>Complications:</u>**<br>•Seizures<br>•Impaired consciousness<br>•Neuropathology: systemic vasculitis (including brain vessels) with neurodegeneration, signs of ischemia. Changes in adenohypophysis. | **<u>Symptoms:</u>**<br>•36% of patients with neurological symptoms.<br>•Neurological symptoms more common in patients with more severe disease courses.<br>•Children: fatigue and headache<br>**<u>Complications:</u>**<br>•(Hypoxic) encephalopathy<br>•Impaired consciousness<br>•Inflammatory PNS/CNS syndromes (including ADEM, GBS and meningitis with leptomeningeal enhancement)<br>•Seizures (conflicting evidence)<br>•Children: encephalopathy<br>•Neuropathology: hypoxic injury, lymphocytic panencephalitis and meningitis, perivenular inflammation |
| **Cross-sectional studies** | •65% neuropsychiatric complaints during the acute and convalescent phase.<br>•Steroid treatment and disease severity predictive for neuropsychiatric symptoms | •Acute necrotizing encephalopathy |
| **Case reports,**<br>**small case series** | •Critical illness neuro-/myopathy (Guillain-Barré syndrome as differential diagnosis)<br>•Seizures<br>•Persistent sleeping difficulties<br>•Delirium<br>•Persistent anosmia<br>•Generalized pain | •Miller-Fisher syndrome<br>•Polyneuritis cranialis<br>•Critical-illness polyneuropathy<br>•Delirium |

*Abbreviations: ADEM = acute demyelinating encephalomyelitis; CNS = central nervous system; CoV = coronavirus; COVID-19 = coronavirus disease 2019; GBS = Guillain-Barré Syndrome; PNS = peripheral nervous system; SARS = severe acute upper respiratory syndrome.*

**Figure 2.**
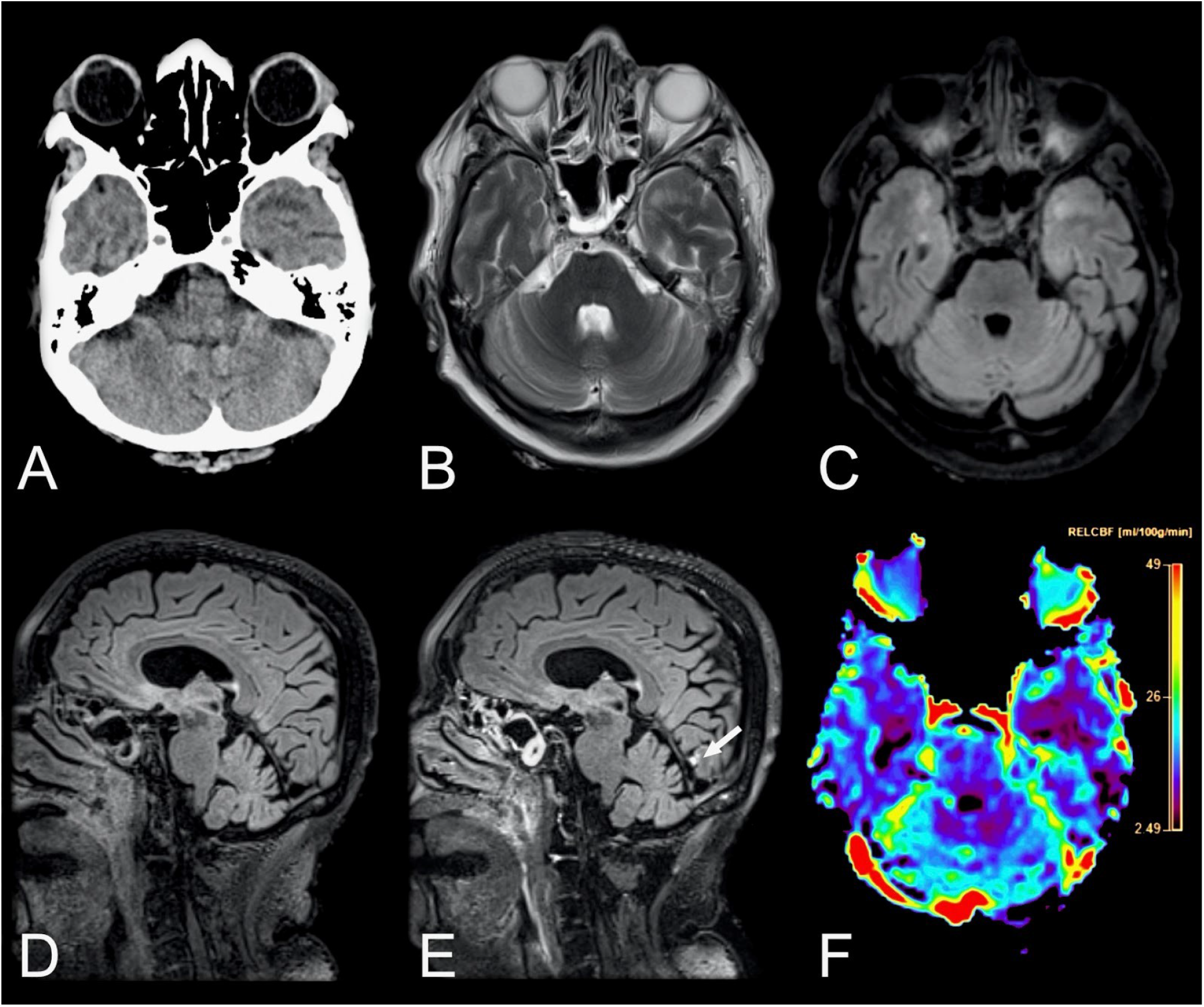
Neuroradiological findings in a 69-year-old female with COVID-19 admitted to the ICU with altered consciousness. Axial non-enhanced head CT scan of the brain (A) showing suspected hypodensities in the temporal poles. Brain MRI was performed two days later. Axial T2-weighted images (B) and T2-weighted FLAIR images (C) confirmed the finding. Pre- and post-contrast sagittal T2- weighted FLAIR images (D, E) in the same patient revealed a solitary focal leptomeningeal enhancement (arrow) by the medial aspect of left occipital lobe, otherwise not easily visible in Ti-weighted contrast-enhanced sequences. The contrast-enhanced perfusion map (F) further revealed reduced blood flow in both temporal lobes. *Abbreviations: COVID-19 = coronavirus disease 2019; FLAIR = fluid attenuated inversion recovery; ICU = intensive care unit*.

## Discussion

The findings of this systematic review suggest that all HCoV strains are associated with neurological manifestations. Specifically, 229E, HKU1, NL63 and OC43 infections have been mainly studied in children, where they are associated with febrile seizures and meningitis. GBS, ADEM and encephalitis have also been described in children. The less abundant evidence for MERS-CoV suggests that fatigue, headache and altered consciousness are common neurological manifestations. Stroke, critical-illness polyneuropathy and GBS have also been observed. Finally, SARS-CoV-1 and SARS-CoV-2, which are genetically highly related,^10^ show substantial similarities regarding the prevalence and presentation of neurological manifestations; symptoms include fatigue, headache and smell/taste disorders. Complications that were observed among both SARS-CoV-1 and SARS-CoV-1 include cerebrovascular pathology, encephalopathy, autoimmune disorders such as GBS, meningoencephalitis and seizures.

### Findings in the context of existing evidence

Overall, HCoV infections show a similar symptom complex across all of the HCoV strains. These symptoms include fatigue and headache. However, shared symptomatology is not surprising since all respiratory virus infections (including rhinoviruses and influenzaviruses) can cause fatigue and headache.^81^ Moreover, these are highly unspecific symptoms that are commonly associated with neurological but also non-neurological diseases.^82^ Thus, these symptoms could arise without direct neuronal involvement of HCoV.

Of note, persistent fatigue has been reported in the recovery/convalescence phase of SARS- CoV-2.^52^ This is reminiscent of persistent fatigue after Epstein-Barr virus (EBV) infection.^83^ Here, the most accepted hypothesis suggests a multifactorial pathogenesis including latent EBV infection, immune and/or neuroendocrine dysfunction as well as behavioral factors. It may be speculated if a similar etiopathogenesis could explain such persistent symptoms also with SARS-CoV-2.

It is evident that SARS-CoV-1 and SARS-CoV-2 not only share neurological symptomatology but also complications. This is not surprising since it has been shown recently that both SARS-CoV-1 and SARS-CoV-2 exploit the same membrane bound molecule angiotensin-converting enzyme 2 as a cellular entry target.^7^ This receptor is, among other tissues, expressed on vessel endothelial cells in the brain, which could serve as a viral entry point to the CNS.^8^ Based on existing literature, five different syndromes can be delineated: (1) cerebrovascular disease, (2) encephalopathy, (3) para-/postinfectious inflammatory CNS/PNS syndromes, (4) (meningoencephalitis and (5) neuropsychiatric complications.

First, ischemic stroke has been reported among both SARS-CoV-1^30^ and SARS-CoV-2 (and anecdotally also for MERS-CoV).^41, 53, 66^ Presumable underlying factors contributing to CNS hypoxia are a prothrombotic state evidenced by high D-dimer levels, which were observed among intensive care unit patients,^40, 42, 84^ disseminated intravascular coagulation (DIC)^85^ and/or systemic vasculitis.^35, 36^ Indeed, widespread endotheliitis has recently been demonstrated in several organs in SARS-CoV-2.^86^ All these conditions facilitate systemic blood clotting. Furthermore, elderly patients, who were commonly reported being affected by stroke, are more likely to have risk factors for cerebrovascular disease.^87^ Additionally, exclusively for SARS-CoV-2, intracranial hemorrhages and cerebral microbleeds have been reported, potentially provoked by endothelial damage/dysfunction.^53, 64, 77^

Second, encephalopathies with clinical features of delirium have been observed among SARS-CoV-2 patients.^53, 66^ Many of these patients showed full clinical recovery.^53^ Anecdotal evidence also indicates encephalopathic changes in SARS-CoV-1.^88^ The underlying mechanisms for the encephalopathy is likely multifactorial, resulting from combinatorial effects of sepsis, hypoxia and immune hyperstimulation (“cytokine storm”).^89^

Third, both studies on SARS-CoV-1 and SARS-CoV-2 reported on GBS, an inflammatory demyelinating disease of the PNS.^65, 90, 91^ Additionally, ADEM, an inflammatory demyelinating CNS disease has been reported among SARS-CoV-2 patients.^53^ Anecdotal evidence also suggests a link of clinically less severe HCoV and MERS-CoV to these diseases. They have a presumable autoimmune pathogenesis^92^ and are thus less likely to be a direct consequence of neuronal infection by HCoV – despite the detection of HCoV RNA in CSF in a few reported cases, but rather represent a para-infectious complication.^90^ Additionally, other viral infections, such as cytomegaly or EBV, have been associated with GBS.^92^ MS is another autoimmune disease whose potential association with HCoV infection has been substantially investigated. The conflicting results do not allow final conclusions on the role of HCoV in MS pathogenesis, even though the majority of evidence contradicts such an association.

Fourth, exclusively for SARS-CoV-2, few patients with clinical and imaging features of (meningo-)encephalitis have been described (in children anecdotally also for HCoV-229E, HKU1, NL63 and/or OC43). These cases were suggestive for primary HCoV CNS infection.^64, 66^ Yet, SARS-CoV-2 RNA in CSF was only detected in a handful of cases.^64, 71^ Fifth, a link between neuropsychiatric complications such as psychosis and mood disorders has been described for SARS-CoV-1, SARS-CoV-2 and clinically more benign HCoV. However, large cohort studies with adequate control groups are needed to address potential confounders, such as secondary consequences of pandemic lockdown measures.

It is noteworthy that SARS-CoV-2 RNA could not be retrieved from the CSF in the vast majority of patients from larger case series/cohort studies.^53, 64, 66^. Similarly, only few incidences of SARS-CoV-1 RNA detection in CSF have been described.^93, 94^ Neuropathological findings support this notion: only one study reported on electron dense particles in CNS neurons suggestive of SARS-CoV-2, though no RNA was detected in the CSF.^80^ Failure to detect SARS-CoV-2 RNA in CSF could depend on different factors: first, the virus could be cell-bound thereby spreading from cell-to-cell; second, virus particles have very low levels in CSF or third, endogenous CSF endo-/exonucleases cleave SARS-CoV-2 RNA.^95^ For the entry of HCoVs into the CNS, two major pathways have been proposed:^96^ the hematogenous (via the blood-brain barrier) and neuronal retrograde pathway (via the cribriform plate/olfactory nerve). The exact pathway is yet still a matter of debate and more studies are necessary to dissect the specific CNS entry mechanisms of HCoVs. However, the absence of SARS-CoV-2 from the CNS might also indicate that a large share of neurological complications are secondary to the severe systemic manifestations of SARS-CoV-2, i.e. from a hyperinflammation syndrome^89^, vasculopathy/coagulopathy, post-infectious auto-inflammation and the effects of sepsis and hypoxia.^53^

### Recommendations for neuroimaging

The results of the current study highlight the need for adequate neuroimaging protocols in HCoV infections with neurological manifestation to be able to accurately detect encephalitis, leptomeningeal enhancement and vascular complications such as microbleeds, subarachnoid hemorrhage and infarcts. Previous studies have mainly relied on CT and MRI without gadolinium-based contrast agents. Cautiousness regarding the use of contrast agents in critically ill patients is naturally warranted, as is the use of ionizing radiation in children. For acute deterioration, a non-enhanced head CT scan is likely the most adequate choice, while more subacute symptomatology or further investigation may necessitate a brain and/or spinal cord MRI. Based on the findings in this review and our emerging experience with SARS-CoV-2, we recommend the MRI protocols suggested in **Table 2** to identify HCoV infection-related neuroradiological manifestations. Examples of neuroimaging findings in SARS-CoV-2 from our clinical practice using the recommended protocols can be found in **Figures 2 and 3**.

**Table 2.**
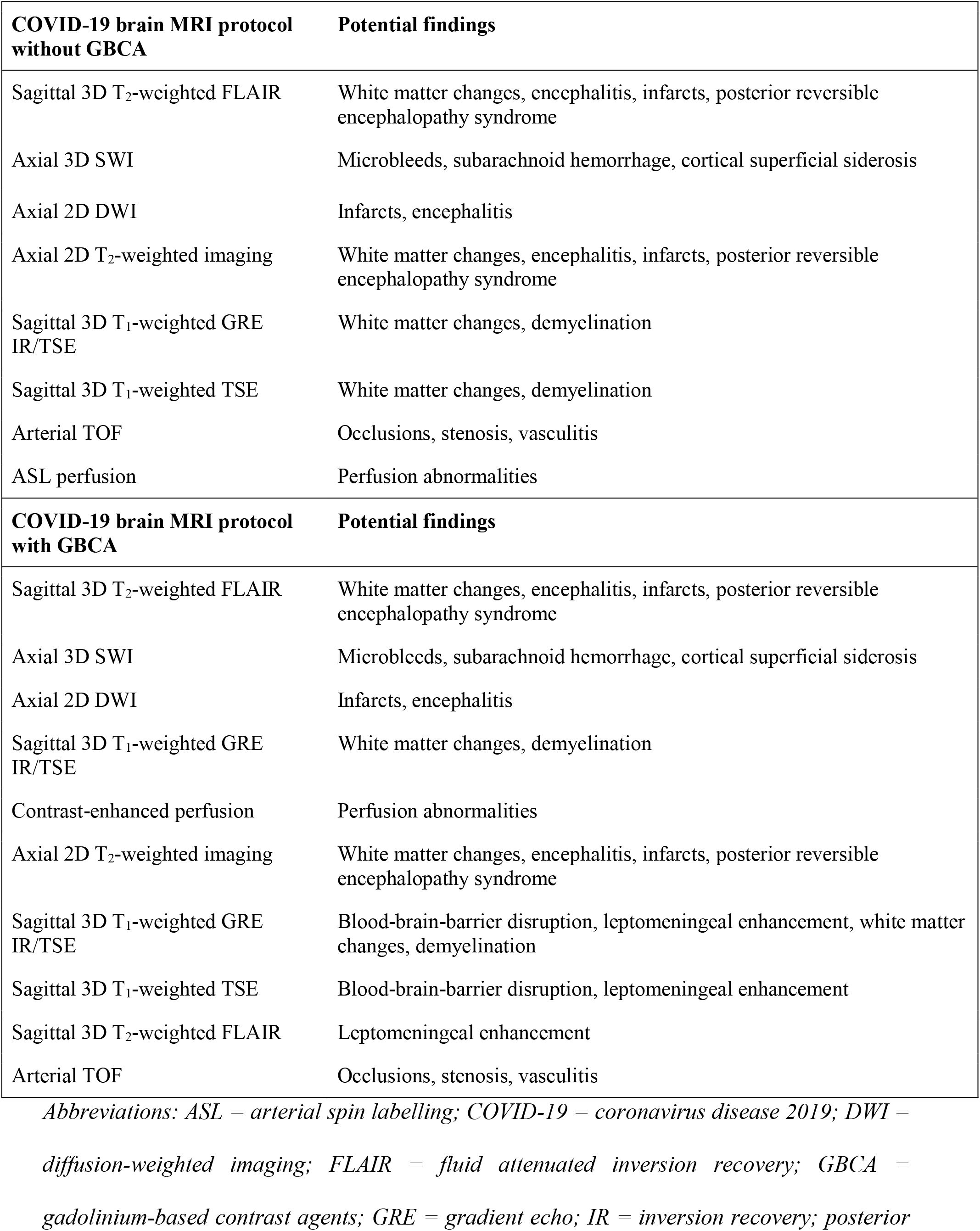
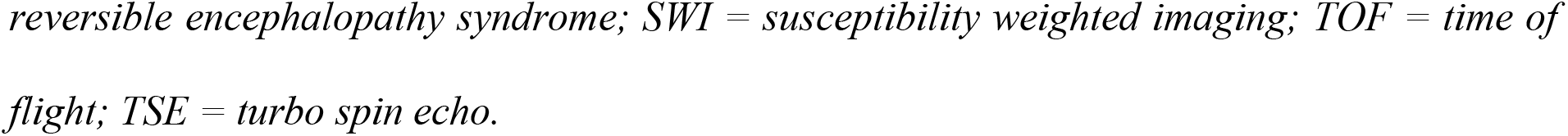
Recommended brain MRI protocol for COVID-19.

| **COVID-19 brain MRI protocol without GBCA** | **Potential findings** |
| --- | --- |
| Sagittal 3D T<sub>2</sub>-weighted FLAIR | White matter changes, encephalitis, infarcts, posterior reversible encephalopathy syndrome |
| Axial 3D SWI | Microbleeds, subarachnoid hemorrhage, cortical superficial siderosis |
| Axial 2D DWI | Infarcts, encephalitis |
| Axial 2D T<sub>2</sub>-weighted imaging | White matter changes, encephalitis, infarcts, posterior reversible encephalopathy syndrome |
| Sagittal 3D T<sub>1</sub>-weighted GRE IR/TSE | White matter changes, demyelination |
| Sagittal 3D T<sub>1</sub>-weighted TSE | White matter changes, demyelination |
| Arterial TOF | Occlusions, stenosis, vasculitis |
| ASL perfusion | Perfusion abnormalities |
| **COVID-19 brain MRI protocol with GBCA** | **Potential findings** |
| Sagittal 3D T<sub>2</sub>-weighted FLAIR | White matter changes, encephalitis, infarcts, posterior reversible encephalopathy syndrome |
| Axial 3D SWI | Microbleeds, subarachnoid hemorrhage, cortical superficial siderosis |
| Axial 2D DWI | Infarcts, encephalitis |
| Sagittal 3D T<sub>1</sub>-weighted GRE IR/TSE | White matter changes, demyelination |
| Contrast-enhanced perfusion | Perfusion abnormalities |
| Axial 2D T<sub>2</sub>-weighted imaging | White matter changes, encephalitis, infarcts, posterior reversible encephalopathy syndrome |
| Sagittal 3D T<sub>1</sub>-weighted GRE IR/TSE | Blood-brain-barrier disruption, leptomeningeal enhancement, white matter changes, demyelination |
| Sagittal 3D T<sub>1</sub>-weighted TSE | Blood-brain-barrier disruption, leptomeningeal enhancement |
| Sagittal 3D T<sub>2</sub>-weighted FLAIR | Leptomeningeal enhancement |
| Arterial TOF | Occlusions, stenosis, vasculitis |

*Abbreviations: ASL = arterial spin labelling; COVID-19 = coronavirus disease 2019; DWI = diffusion-weighted imaging; FLAIR = fluid attenuated inversion recovery; GBCA = gadolinium-based contrast agents; GRE = gradient echo; IR = inversion recovery; posterior*
*reversible encephalopathy syndrome; SWI = susceptibility weighted imaging; TOF = time of flight; TSE = turbo spin echo.*

**Figure 3.**
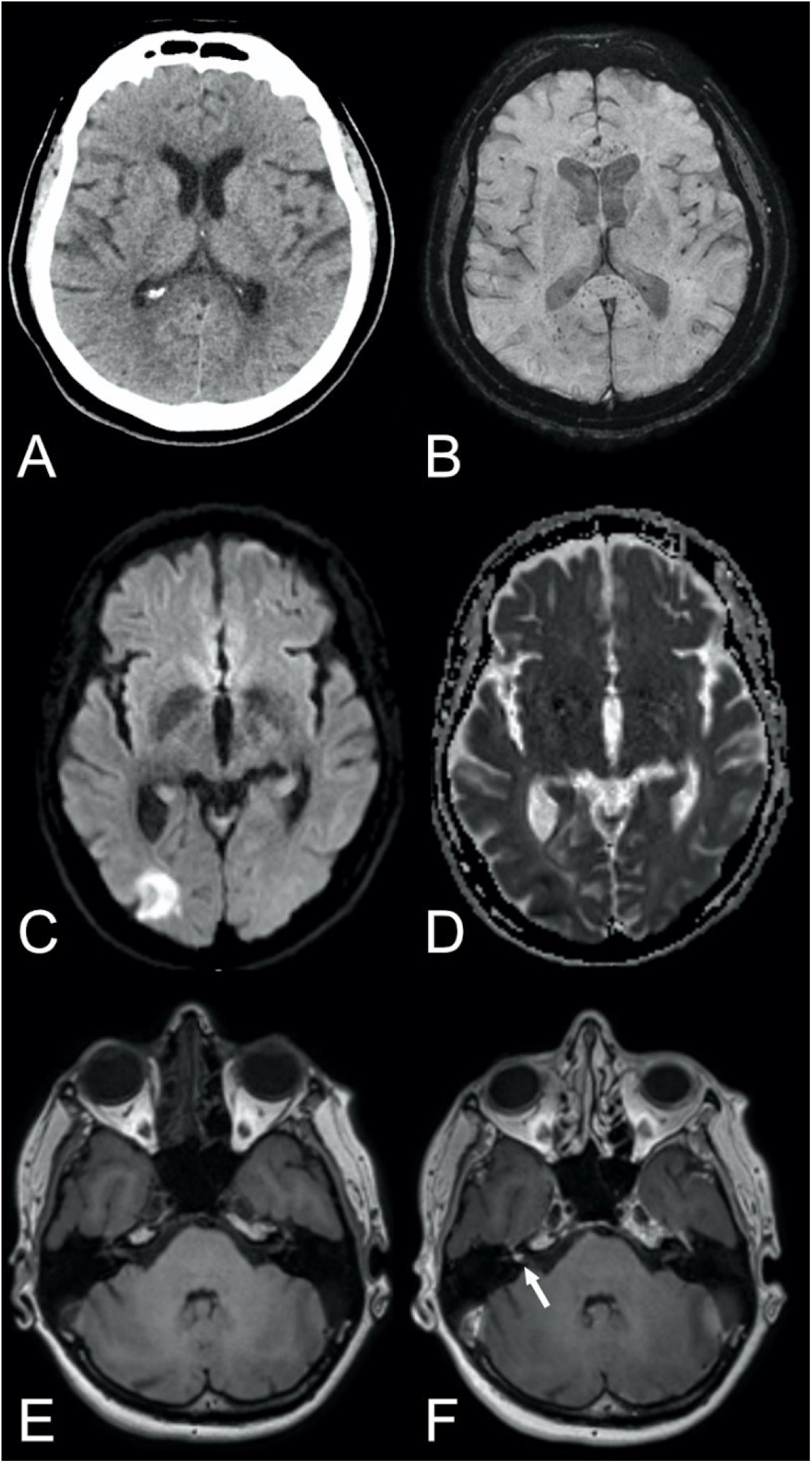
Various neuroradiological findings in COVID-19 patients. ***First row:*** Axial non-enhanced head CT, reported as normal (A) but brain MRI, including susceptibility-weighted imaging (B) revealed numerous microbleeds with a predilection to the corpus callosum and especially the splenium of corpus callosum. The aforementioned case exemplifies the importance of susceptibility-weighted imaging in the neuroimaging of COVID-19 patients in means of MRI brain examination. ***Second row:*** Axial diffusion-weighted imaging, b1000 (C) and ADC map (D) showing a cortical infarct in the right occipital lobe. ***Third row:*** Axial T_1_-weighted images pre- and post contrast (E, F) depicting a focal enhancement of the right vestibular nerve. *Abbreviations: ADC = apparent diffusion coefficient; COVID-19 = coronavirus disease 2019*.

### Limitations

Due to potential delay in publishing relevant findings, a significant proportion of evidence on SARS-CoV-2 may yet to be published. Nevertheless, the available evidence suggests striking similarities of neurological manifestations between SARS-CoV-1 and SARS-CoV-2 patients.

### Conclusions

Our systematic review provides high level evidence that HCoVs, particularly SARS-CoV-1 and SARS-CoV-2, have a common ground on neurological symptomatology and complications. Particularly, clinicians treating these patients should be vigilant for at least five classes of neurological complications: (1) Cerebrovascular disorders including ischemic stroke and macro/micro-hemorrhages, (2) encephalopathies, likely caused by combinatorial effects of sepsis, hypoxia and immune hyperstimulation, (3) para-/postinfectious immune-mediated complications such as GBS and ADEM, (4) (meningo-)encephalitis, potentially with concomitant seizures and (5) neuropsychiatric complications such as mood disorders or psychosis. Thus, the clinical neuroimaging protocol, as herein recommended, should include appropriate MRI sequences for early diagnosis and monitoring of patients. Our results also underscore the need to further research and optimize treatment regarding modulation of the coagulation and immune systems in order to reduce the risk of neurological complications.

## Data Availability

Data is available upon request to the corresponding author.

## Acknowledgements

We thank Francis Poulenc for help with data analysis and Ioannis Koupidis for help with figure panels.

## Author contribution

Jesper Almqvist: Designed and conceptualized the study, data acquisition, analysis and interpretation, drafting and revision of the manuscript; Tobias Granberg: Designed and conceptualized the study, data acquisition, analysis and interpretation, drafting and revision of the manuscript; Antonios Tzortzakakis: Data interpretation, revision of the manuscript for intellectual content; Stefanos Klironomos: Data interpretation, revision of the manuscript for intellectual content; Evangelia Kollia: Data interpretation, revision of the manuscript for intellectual content; Claes Öhberg: Data interpretation, revision of the manuscript for intellectual content; Roland Martin: Data interpretation, revision of the manuscript for intellectual content; Fredrik Piehl: Data interpretation, revision of the manuscript for intellectual content; Russell Ouellette: Data interpretation, revision of the manuscript for intellectual content; Benjamin V. Ineichen: Designed and conceptualized the study, data acquisition, analysis and interpretation, drafting of the manuscript

## Ethical approval

Reporting figures with examples from our own practice was approved by the Swedish Ethical Review Authority; informed consent was waived due to the retrospective nature of the retrospective case reports.

## Supplementary files

Supplementary tables e-1 to e-8:

e-1: Search string

e-2: Pre-defined criteria Newcastle-Ottawa scale for risk of bias e-3: Risk of bias assessment

e-4: Summary of studies assessing neurological manifestations and complication in the more clinically mild HCoVs: HCoV-229E, HKU1, NL63 and/or OC43 infections.

e-5: Summary of studies assessing neurological manifestations and complication in MERS-CoV infections.

e-6: Summary of studies assessing neurological manifestations and complication in SARS-CoV-1 infections

e-7: Summary of studies assessing neurological manifestations and complication in SARS-CoV-2 infections.

e-8: Association of coronaviruses with multiple sclerosis.

Supplementary reference list

## Glossary

ADEM = acute disseminated encephalomyelitis, ANE = acute necrotizing encephalopathy, COVID-19 = coronavirus disease 2019, CSF = cerebrospinal fluid, CT = computed tomography, DIC = disseminated intravascular coagulation, EEG = electroencephalogram, GBS = Guillain-Barre syndrome, HCoV = human coronavirus, MERS = Middle East respiratory syndrome, MRI = magnetic resonance imaging, MS = multiple sclerosis, PNS = peripheral nervous system, PRES = Posterior reversible encephalopathy syndrome, PRISMA = Preferred Reporting Items for Systematic Reviews and Meta-Analysis, SARS = severe acute respiratory syndrome

## Journal

Annals of Clinical and Translational Neurology

